# Excess of Cardiovascular Deaths During the COVID-19 Pandemic in Brazilian Capital Cities

**DOI:** 10.1101/2020.06.24.20139295

**Authors:** Luisa C. C. Brant, Bruno R. Nascimento, Renato Teixeira, Marcelo Antônio C. Q. Lopes, Deborah C. Malta, Gláucia Maria M. Oliveira, Antonio Luiz P. Ribeiro

**Affiliations:** Faculdade de Medicina, Universidade Federal de Minas Gerais, Belo Horizonte, MG, Brazil; Hospital das Clínicas, Universidade Federal de Minas Gerais, Belo Horizonte, MG, Brazil; Sociedade Brasileira de Cardiologia, Rio de Janeiro, RJ, Brazil; Hospital Alberto Urquiza Wanderley, João Pessoa, PB – Brasil; Escola de Enfermagem, Universidade Federal de Minas Gerais, Belo Horizonte, MG, Brazil; Faculdade de Medicina, Universidade Federal do Rio de Janeiro, Rio de Janeiro, RJ, Brazil; Instituto do Coração Edson Saad, Universidade Federal do Rio de Janeiro, Rio de Janeiro, RJ, Brazil

**Keywords:** COVID-19, mortality, cardiovascular disease, acute coronary syndrome, Brazil

## Abstract

**Introduction:** During the COVID-19 pandemic, excess mortality has been reported, while hospitalizations for acute cardiovascular events reduced. Brazil is the second country with more deaths due to COVID-19. We aimed to evaluate excess cardiovascular mortality during COVID-19 pandemic in 6 Brazilian capital cities.

**Methods:** Using the Civil Registry public database, we evaluated total and cardiovascular excess deaths, further stratified in ACS, stroke and unspecified cardiovascular deaths in the 6 Brazilian cities with greater number of COVID-19 deaths (São Paulo, Rio de Janeiro, Fortaleza, Recife, Belém, Manaus). We compared data from epidemiological weeks 12 to 22 of 2020, with the same period in 2019. We also compared the number of hospital and home deaths during the period.

**Results:** There were 69,328 deaths and 17,877 COVID-19 deaths in the studied period and cities for 2020. Cardiovascular mortality increased in most cities, with greater magnitude in the Northern capitals. However, while there was a reduction in ACS and stroke in the most developed cities, the Northern capitals showed an increase of these events. For unspecified cardiovascular deaths, there was a marked increase in all cities, which strongly correlated to the rise in home deaths (r=0.86, p=0.01).

**Conclusion:** The excess cardiovascular mortality was greater in the less developed cities, possibly associated with healthcare collapse. ACS and stroke deaths decreased in the most developed cities, in parallel with an increase in unspecified cardiovascular and home deaths, presumably as a result of misdiagnosis. Conversely, ACS and stroke deaths increased in cities with a healthcare collapse.

**Clinical Perspective:**

- **What is already known about this subject?** During the pandemic, beyond deaths due to confirmed COVID-19, there seems to be an increase in the total number of deaths compared to previous years in Brazil. Excess mortality may have occurred due to identified or not COVID-19 or other causes, being an objective and comparable metric for healthcare evaluation.
- **What does this study add?** In the 6 Brazilian capitals with higher numbers of deaths due to COVID-19, the impact of the pandemic in the excess all-cause and cardiovascular deaths was noticeable, especially in regions where health systems collapsed, which are the most socioeconomically deprived. In the other capital cities, the decreasing number of deaths associated with well-defined events (ACS and stroke) paralleled with more frequent undefined cardiovascular and home deaths.
- **How might this impact on clinical practice?** Investments should be prioritized to areas where the pandemic resulted in health system collapse. During periods of social distancing, campaigns and strategies to increase the population’s awareness of cardiovascular care, health promotion practices, seeking services in the case of acute signs and symptoms, should be prioritized by governments. The Corresponding Author has the right to grant on behalf of all authors and does grant on behalf of all authors, an exclusive license on a worldwide basis to the BMJ Publishing Group Ltd and its Licensees to permit this article to be published in HEART editions and any other BMJPGL products to exploit all subsidiary rights.

## Introduction

The COVID-19 pandemic was decreed by the World Health Organization on March 11^th^, 2020, pointing towards a great potential for global spread and significant mortality rates. Since the first case was notified in Brazil in February 26^th^, the epidemic has evolved rapidly, and in June 23^th^, more than 1,136,470 cases have been reported in the country, with almost 51,271 deaths^1^.

Beyond deaths due to confirmed COVID-19, previous reports have also emphasized an increase in the number of total deaths during the pandemic compared to the same period of previous years: the excess mortality. From March to May, 2020, the excess of deaths in Italy was 49%, reaching 277% in New York City^2^. Of note, excess deaths in this period may have occurred due to identified or non-identified COVID-19 or other causes, being an objective and comparable metric for evaluating the true impact of the pandemic in the mortality of a location^3^.

Concomitantly, there has been a decrease in hospital admissions associated with acute cardiovascular diseases, markedly acute coronary syndromes (ACS), in high-income countries (HIC). In Northern Italy, a significant decrease in hospital daily admissions due to ACS during the pandemic was reported^4^. In parallel, a 58% increase in out-of-hospital cardiac arrest, strongly associated with cumulative COVID-19 incidence, was also observed^5^. In the US, an estimated 38% reduction in cardiac catheterization laboratories activations due to ST-elevation myocardial infarction (STEMI) was described^6^, similar to the 40% reduction in Spain^7^. This behavior may be associated with avoidance of medical care due to social distancing, concerns of contracting COVID-19 in the hospital, and misdiagnosis. Moreover, in regions were healthcare resources became scarce during the pandemic, excess deaths may have occurred due to healthcare collapse, reinforcing social disparities in the death tolls.

The pandemic has been delayed in low- and middle-income countries (LMIC), and how cardiovascular diseases will behave in LMIC is a matter of debate. Brazil, a middle-income country, is ranked second in the number of deaths due to COVID-19^1^. However, deaths are heterogeneously distributed across the country and numbers are certainly underestimated due to the low rate of COVID-19 diagnostic tests performed^1^. Using data from the civil registry database, we aimed to evaluate excess cardiovascular mortality during the COVID-19 pandemic in 6 Brazilian capital cities from March 15^th^ to May 22^nd^, 2020.

## Methods

### Data Sources

Data analytic methods and study materials will be made available to other researchers for purposes of reproducing the results or replicating the procedure, from the corresponding author upon reasonable request.

We conducted an observational retrospective study using the Civil Registry public database (Transparency Portal)^8^ from ARPEN-Brazil for mortality data. Founded in September 1993, ARPEN (*Associação dos Registradores de Pessoas Naturais*, National Association of Natural Persons Registrars) represents the class of Civil Registry Officers in the country, serving all states, and performing the main acts and certifications of civil life, including death certificates (DC). We used registers from the 6 Brazilian capitals with more COVID-19 deaths: São Paulo, Rio de Janeiro, Fortaleza, Recife, Belém, and Manaus, from the epidemiological week of the first documented death due to COVID-19 in Brazil (March 17^th^) to May 22^nd^ 2020 (12^th^ to the 22^nd^ epidemiological weeks), and compared to the same period in 2019. Data was not compared to the Brazilian Mortality Information System (SIM), which is officially used by the Brazilian government, because it is usually only publicly available in the following year.

**Table 1** shows the region of Brazil, population, health resources, and human development index (HDI) for each of the 6 capital cities. The HDI index for each capital city was extracted from *Atlas de Desenvolvimento Humano no Brasil*^*9*^, being a summary measure of socioeconomic development, which uses the indicators: life expectancy, years of schooling, and gross national income per capita; and is expressed on a scale of 0 to 1, being better if closer to 1^10^. The number of intensive care unit (ICU) beds of each capital cities was extracted from the most updated Geography and Statistics Brazilian Institute (*Instituto Brasileiro de Geografia e Estatística* – IBGE) database, and the numbers of physicians and nurses per 100,000 inhabitants derived from the Public Health System database (DataSUS)^11^. These variables were used as proxies for healthcare resources.

**Table 1:** Sociodemographic characteristics and total deaths in 6 selected Brazilian capital cities during epidemiological weeks 12 to 22, in 2019 and 2020.

|  | São Paulo | Rio de Janeiro | Fortaleza | Recife | Belém | Manaus |
| --- | --- | --- | --- | --- | --- | --- |
| *Region* | *Southeast* | *Southeast* | *Northeast* | *Northeast* | *North* | *North* |
| Population, 2019 | 12,097,289 | 6,587,583 | 2,639,652 | 1,604,920 | 1,370,685 | 2,180,647 |
| Population, 2020 | 12,142,621 | 6,592,228 | 2,651,800 | 1,607,059 | 1,359,971 | 2,216,197 |
| HDI | 0.81 | 0.80 | 0.75 | 0.77 | 0.75 | 0.74 |
| ICU beds per 100,000 | 28.6 | 38.5 | 21.9 | 60.3 | 30.2 | 13.3 |
| Physicians per 100,00 | 353 | 329 | 251 | 407 | 234 | 167 |
| Nurses per 100,000 | 192 | 192 | 171 | 268 | 114 | 117 |
| Total Deaths, 2019 | 19,593 | 14,032 | 4,629 | 3,472 | 2,285 | 2,552 |
| Total Deaths Rates, 2019 | 162 | 213 | 175 | 216 | 167 | 117 |
| Total Deaths, 2020 | 25,426 | 19,180 | 8,345 | 5,745 | 4,899 | 5,733 |
| Total Deaths Rates, 2020 | 209 | 291 | 315 | 358 | 360 | 259 |
| Excess deaths | 5,833 | 5,148 | 3,716 | 2,273 | 2,614 | 3,181 |
| Excess deaths (%) | 30 | 37 | 80 | 66 | 114 | 125 |
| COVID-19 deaths | 5,343 | 5,277 | 3,623 | 1,119 | 1,399 | 1,116 |
| Excess - COVID-19 deaths | 490 | -129 | 93 | 1154 | 1215 | 2065 |
| Excess - COVID-19 deaths (%) | 3 | -1 | 2 | 33 | 53 | 81 |

Abbreviations: ICU: intensive care unit, HDI: human development index.

Our data are based on the DC registered in Notaries, with only one cause being presented for each death. In a regular DC, the cause(s) of death is(are) declared in Part I (lines a, b, c, d), with the underlying cause in the last line, according to the ICD-10 rules. Part II records other significant causes that may have contributed to death. In this analysis of the Transparency Portal database, in addition to COVID-19 reported as a suspected or confirmed cause, we also assessed other causes related to the new coronavirus disease, shown in **Table 2**.

**Table 2:** Hierarchical data mining procedure to assess natural causes of death in the death certificate to define one cause per death, and other related conditions.

| Condition | Hierarchical definition |
| --- | --- |
| **Condition 1** | Mention of COVID-19, coronavirus: considered as “COVID-19” (suspected or confirmed); |
| ○ **Condition 1.1** | • Mention of ACS / infarction, associated with COVID-19, coronavirus: considered as “ACS with COVID-19”; |
| ○ **Condition 1.2** | • Mention of stroke (ischemic or hemorrhagic ), associated with COVID-19: considered as “Stroke with COVID-19”; |
| **Condition 2** | Mention of SARS: considered as “SARS”; |
| **Condition 3** | Mention of ACS / infarction not associated with COVID-19, coronavirus: considered as “ACS without COVID-19” (ACS); |
| **Condition 4** | Mention of stroke (ischemic or hemorrhagic) not associated with COVID-19, considered “stroke without COVID-19” (Stroke); |
| **Condition 5** | Mention of Pneumonia associated with non-cardiovascular causes (excluding those listed above): considered as “Pneumonia”; |
| **Condition 6** | Mention of an undetermined cause, sudden death or cardiorespiratory arrest, associated with arterial hypertension, diabetes mellitus, pulmonary embolism, heart failure, dilated cardiomyopathy, pulmonary edema, atrioventricular block, cardiac arrhythmia, supraventricular tachycardia, ventricular tachycardia, fibrillation atrial, bradyarrhythmia: considered “unreported cause associated with cardiovascular disease”; |
| **Condition 7** | Mention of "Sudden death": considered "Sudden death"; |
| **Condition 8** | Mention of cardiogenic shock, associated with ischemic disease, was considered “cardiogenic shock associated with ischemic disease”; |
| **Condition 9** | Mention of Sepsis as the only reported cause: considered as “Sepsis”; |
| **Condition 10** | Mention of Respiratory failure as the only reported cause: considered as “Respiratory failure”; |
| **Condition 11** | Mention of Indeterminate Cause as the only reported cause: considered as “Indeterminate cause”; |
| **Condition 12** | Death not classified under any of the previous conditions: considered as "Other Cause". |

**Abbreviations:** ACS: acute coronary syndrome; SARS: severe acute respiratory syndrome.

In this analysis, we classified deaths associated with conditions reported in **Table 2** according to: a) date of death, per day and month in 2019 and 2020; b) location (Brazil, by states and cities), considering the place of death declared in the DC; and c) place of death (hospital or home). The Transparency Portal^8^ is frequently updated by the Civil Registry Offices following legal deadlines: the family has up to 24 hours to register the death in the notary, which has up to 5 days to perform the official registration, and up to 10 days to send the final act to the National Civil Registry Information Center, which updates the public database. Of note, data is only publicly available for 2019 and 2020, precluding comparison with previous years.

### Patient involvement

Patients and public were not involved in the design and conduct of this research.

### Statistical Analysis

A hierarchical procedure was applied to the raw database^8^ on June 19^th^, 2020, and data was mined according to the dictionary of search terms detailed in **Supplemental Table 1**. The search terms were grouped based on a standardized dictionary and treated based on the IRIS software^12^ and text mining methods were additionally applied to search for terms that identify the final diseases. A hierarchical procedure was then carried out to assess all natural (non-external) causes declared in the DC and select only 1 cause per death, and other related conditions, as detailed in **Table 2**.

For the purpose of this study, the following variables regarding mortality were used for each city: total number of deaths in 2019 and 2020; excess mortality calculated by subtracting the observed numbers of deaths in the study period from the deaths that occurred during the same period in 2019; and excess mortality, excluding COVID-19 deaths. Moreover, we report number of deaths and excess mortality for ACS (condition 3), stroke (condition 4), unspecified cardiovascular disease (Sum of conditions 6, 7 and 8), and total cardiovascular deaths (sum of conditions 3, 4, 6, 7 and 8). Numbers and percent changes were reported. Moreover, deaths were classified by place of death (hospital and home). Mortality rates per 100,000 inhabitants were calculated dividing the number of deaths of a specific condition by the estimated population of each capital in the same year^13 14^. Mortality rates herein reported are not age-standardized.

Lastly, we evaluated the Pearson correlation coefficient for unspecified cardiovascular deaths and home deaths. A bicaudal p-value <0.05 was considered statistically significant.

## Results

**Table 1** shows demographic, health resources data and the number of total and excess all-cause deaths in the 6 selected capital cities. Capital cities in the Southeast region (São Paulo and Rio de Janeiro) are more developed, compared to cities in the Northeast (Fortaleza and Recife) and North (Belém and Manaus) regions, as denoted by the HDI. Comparing to the same period in 2019, there was an excess in total deaths in 2020 for all capital cities, with greater magnitude in the cities with lower HDI. However, when deaths due to COVID-19 were excluded from excess deaths, excess mortality was only relevant in the capital cities in the North region and Recife. **Figure 1** depicts that the excess deaths in São Paulo, Rio de Janeiro and Fortaleza are mostly explained by COVID-19 deaths, while in Belém and Manaus, there was an excess due to other causes. In Recife, there was a high number of SARS deaths what, together with COVID-19 deaths, explained the excess mortality. As such, excess mortality by causes other than COVID-19 and SARS were only relevant in Belém and Manaus (North).

**Figure 1:**
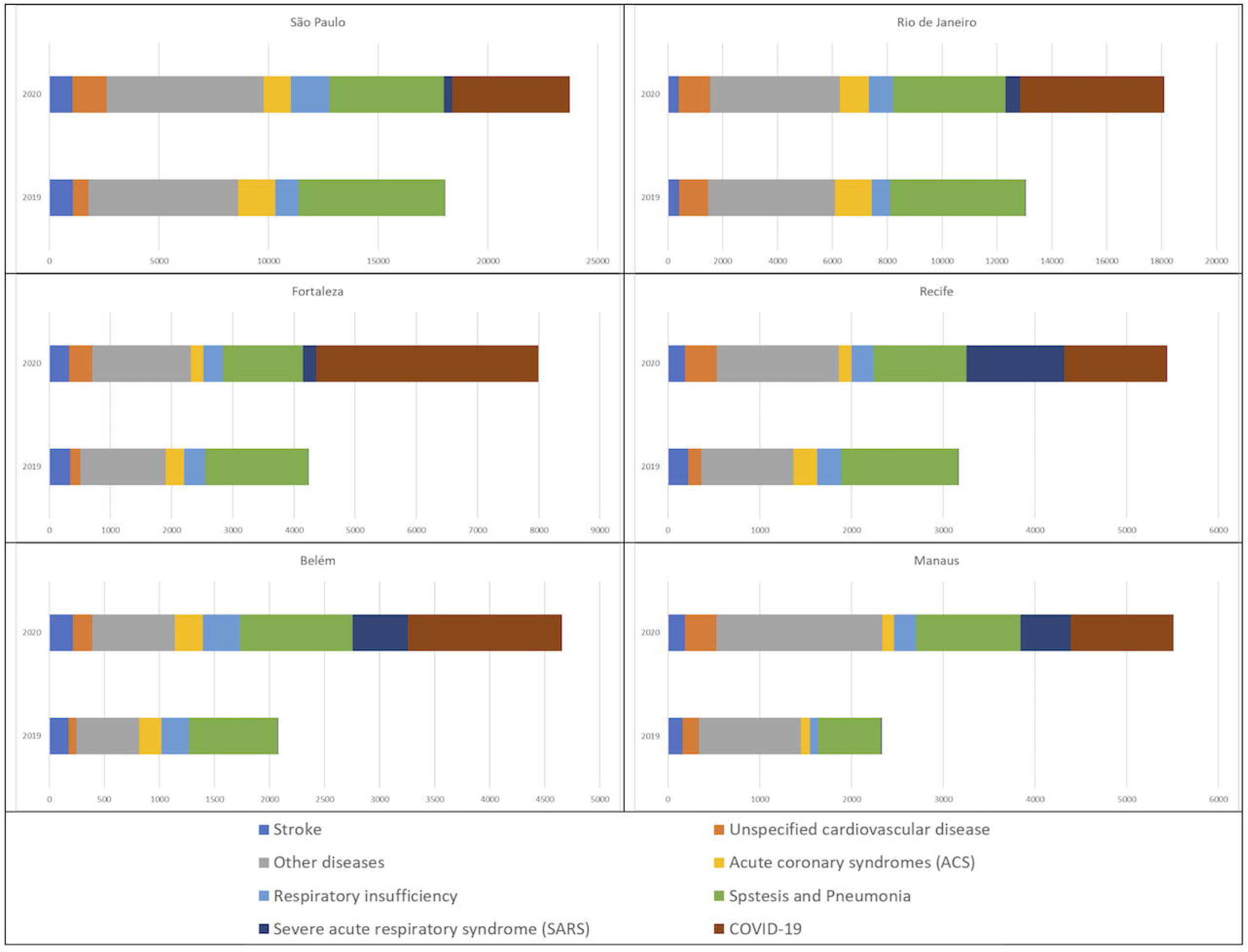
Horizontal bar graphs with mortality per pre-defined causes (from the data mining algorithm, including cardiovascular causes), per selected Brazilian capital city, in 2019 and 2020.

Regarding cardiovascular deaths, **Table 3** reveals that, except for Rio de Janeiro, there was an excess in total cardiovascular deaths in all cities, again with greater magnitude in the Northern cities. When considering specific causes, there was a reduction in ACS deaths, except in the Northern capitals, where there was an increase. The same pattern described for ACS can be seen for stroke, however with lesser magnitude. For unspecified cardiovascular deaths, there was a marked increase in all capital cities. **Supplement Figure 1** shows these changes along the studied period, revealing that the rise in unspecified cardiovascular deaths occurred in parallel with the reduction of ACS. **Figure 2** illustrates the above data, showing the percent change in deaths per capital city caused by definite cardiovascular causes (ACS and stroke) and unspecified cardiovascular causes.

**Table 3:** Cardiovascular, acute coronary syndromes, stroke and unspecified cardiovascular deaths in 6 selected Brazilian capital cities during epidemiological weeks 12 to 22, in 2019 and 2020.

|  | **São Paulo** | **Rio de Janeiro** | **Fortaleza** | **Recife** | **Belém** | **Manaus** |
| --- | --- | --- | --- | --- | --- | --- |
| *Region* | *Southeast* | *Southeast* | *Northeast* | *Northeast* | *North* | *North* |
| Total CV deaths, 2019 | 3768 | 2956 | 876 | 682 | 498 | 486 |
| Total CV deaths, 2020 | 4198 | 2810 | 967 | 735 | 705 | 701 |
| Difference in Total CV deaths | 430 | -146 | 91 | 53 | 207 | 215 |
| Difference in Total CV deaths (%) | 11.4 | -4.9 | 10.4 | 7.8 | 41.6 | 44.2 |
| ACS deaths, 2019 | 1830 | 1420 | 329 | 283 | 219 | 108 |
| ACS deaths, 2020 | 1396 | 1163 | 224 | 162 | 278 | 131 |
| Difference in ACS deaths | -434 | -257 | -105 | -121 | 59 | 23 |
| Difference in ACS deaths (%) | -23.7 | -18.1 | -31.9 | -42.8 | 26.9 | 21.3 |
| Stroke Deaths, 2019 | 1193 | 442 | 363 | 240 | 193 | 178 |
| Stroke Deaths, 2020 | 1180 | 419 | 353 | 212 | 239 | 207 |
| Difference in Stroke deaths | -13 | -23 | -10 | -28 | 46 | 29 |
| Difference in Stroke deaths (%) | -1.1 | -5.2 | -2.8 | -11.7 | 23.8 | 16.3 |
| Unspecified CV deaths, 2019 | 745 | 1094 | 184 | 159 | 86 | 200 |
| Unspecified CV deaths, 2020 | 1622 | 1228 | 390 | 361 | 188 | 363 |
| Difference in Unspecified CV deaths | 877 | 134 | 206 | 202 | 102 | 163 |
| Difference in Unspecified CV deaths (%) | 117.7 | 12.2 | 112.0 | 127.0 | 118.6 | 81.5 |

**Abbreviations:** ACS: acute coronary syndrome; CV: cardiovascular.

**Figure 2:**
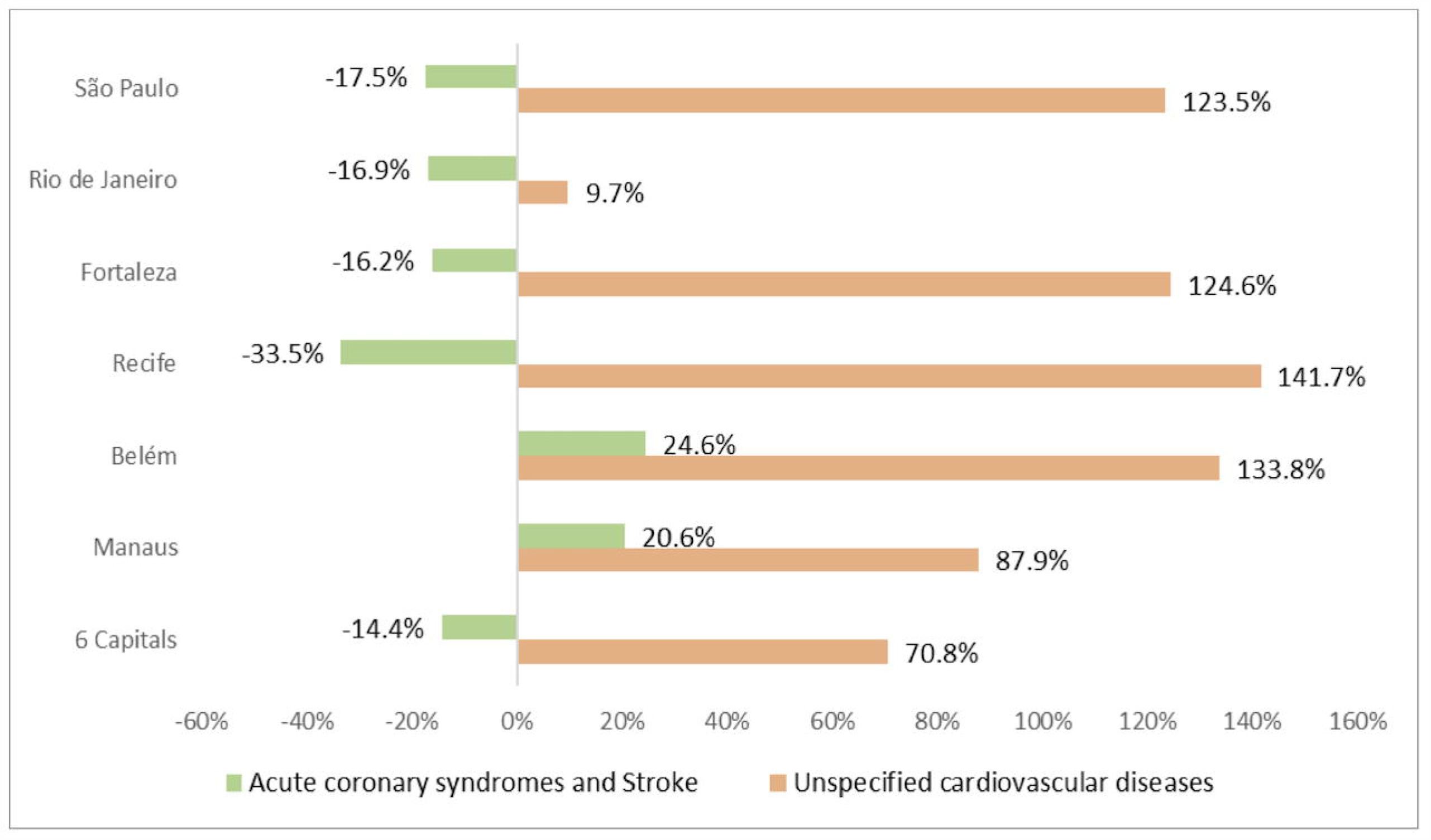
Percent change in the number of deaths between 2019 and 2020 for acute coronary syndromes and unspecified cardiovascular diseases per selected 6 capital cities.

Regarding place of death, **Figure 3A** and **3B** demonstrates the number of hospital and home deaths along the studied period in 2020 compared to 2019. Although the increase in deaths is seen in both settings, in hospitals it was mainly explained by COVID-19 and SARS, while the same did not happen for home deaths.

**Figure 3:**
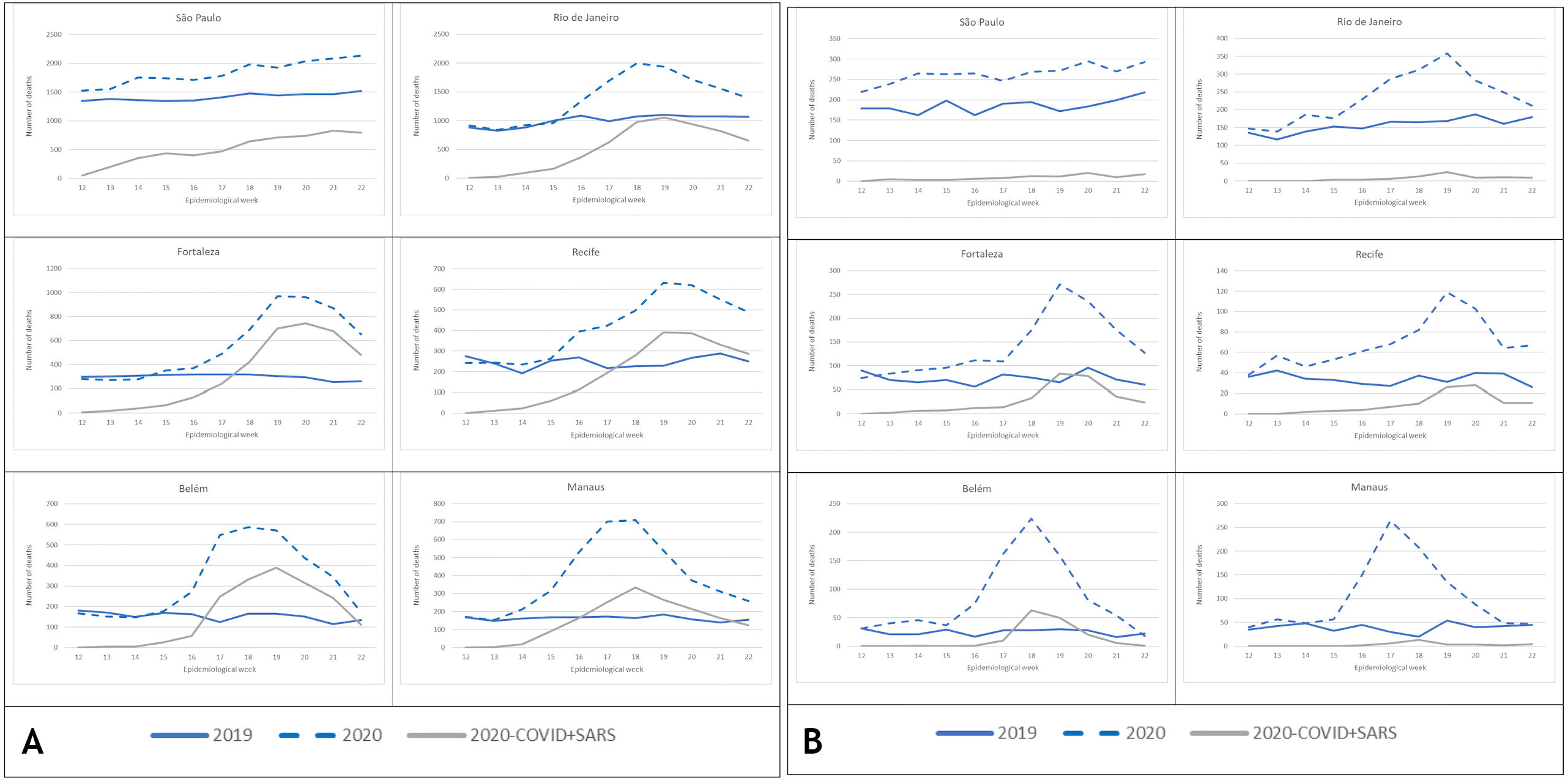
**A:** Total number of hospital deaths in 2019 and 2020 and deaths associated with COVID-19 and severe acute respiratory syndrome in 2020. **B:** Total number of home deaths in 2019 and 2020 and deaths associated with COVID-19 and severe acute respiratory syndrome in 2020.

Lastly, **Figure 4** shows that the increase in unspecified cardiovascular deaths in 2020 during the studied period had a positive and strong correlation to the number of home deaths (r=0.86, p=0.01).

**Figure 4:**
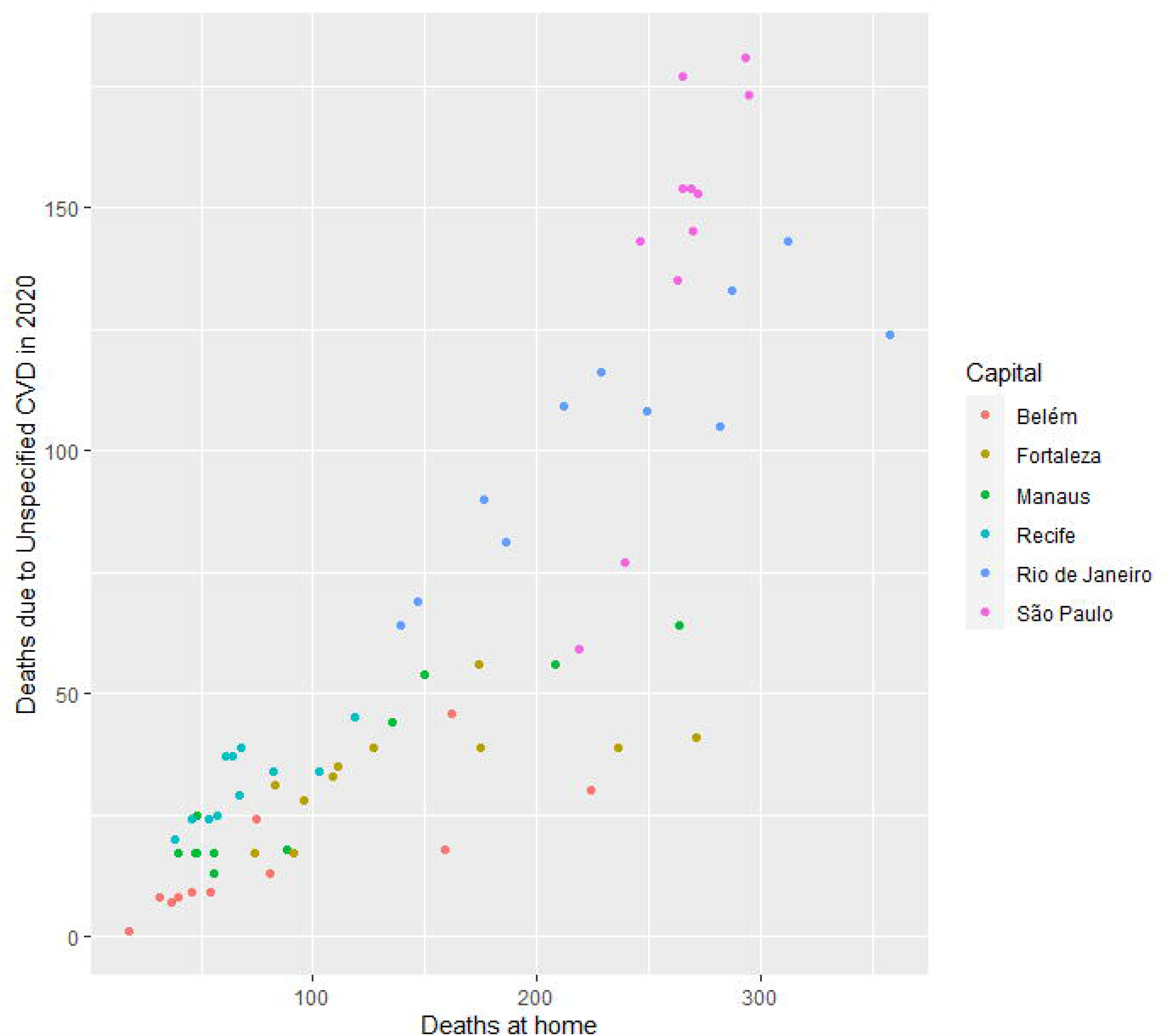
Correlation between home deaths and deaths from unspecified cardiovascular causes in 6 selected Brazilian capital cities in 2020.

## Discussion

Our data – the first comprehensive analysis of the Brazilian civil registry looking for specific patterns of excess mortality during the COVID-19 pandemic – show an excess of total mortality in the 6 capital cities with highest numbers of COVID-19 deaths. However, while in the Southeastern and Northeastern capital cities COVID-19 and SARS explained most of the total excess mortality, the pattern differed in Northern capitals, where there was a marked increase in all analyzed causes. Excess cardiovascular mortality occurred in most cities, due to a rise in unspecified cardiovascular causes, except in the Northern cities, where ACS and stroke also increased significantly. In the other cities, there was a reduction of deaths due to ACS and stroke, which paralleled with increasing numbers of unspecified cardiovascular deaths, strongly correlated to the rise of home deaths. The increase in ACS and stroke deaths in Northern cities differ from what has been described in HIC, and may anticipate the behavior of ACS and stroke deaths in deprived regions, prone to collapse of health systems.

The Brazilian civil registry database has the advantage of being the only promptly available data source for mortality, with relatively short delays, important features during a pandemic. On the other hand, the records serve demographic – and not epidemiological – purposes, and are not the official source of mortality data for Brazil. Thus, there are no investigation procedures, codification or redistribution of causes of deaths. As such, the quality of the data depends on how correctly the death certificates are filled out, which can differ between locations, as can delays. As such, the official SIM data may diverge in the future, in relation to the Civil Registry. To minimize the delays in notifications, we opted to include only capital cities in the present analysis. Furthermore, as we aimed to investigate the impact of COVID-19 on excess mortality, the choice for locations with highest numbers of deaths from the disease (above 1,000) may provide better insights about health system overload and effects on deaths from other causes.

There are several issues in the distribution of health resources in Brazil, which are concentrated in the more developed locations^15^. From the cities included in this analysis, it’s possible to infer a clear gradient between HDI – a measure of socioeconomic development – and excess mortality, especially in the 2 Northern cities of the lower socioeconomic bound. This is possibly associated with the baseline preparedness of local health systems, regarding hospital infrastructure – especially in the tertiary and quaternary levels – and availability of high-level staff^16^, as well as access to healthcare and emergency transport systems. Medical workforce is unequally distributed in Brazil, and the lowest numbers of nurses, physicians and specialists per inhabitant are in the North region^16 17^. Interestingly, the number of ICU beds per 100,000 inhabitants was not a proxy for healthcare status in this analysis, although collapse of intensive care resources was reported as a marker of disastrous epidemiological scenarios^18 19^. This finding may be explained by the fact that healthcare resources in less developed Brazilian states, particularly in the North region, is concentrated in the capital cities, which therefore suffer the impact of the pandemic in the whole state^11^. Furthermore, the territories of the Amazonas and Pará states, in the Amazon region, are vast and lack paved roads and transport infrastructure, what certainly potentialized the impacts of COVID-19, markedly on home and unspecified events. This suggests that the relation of socioeconomic conditions and excess mortality is beyond the disparity in health resources^20^.

Besides local infrastructure, the availability of COVID-19 tests differed widely between Brazilian regions – even within the same region^1^ – and this might have contributed to the heterogeneity of death reports. Presumably, there was underreporting in the whole country, considering the estimated number of tests per million inhabitants^1^. Also, the cities included in this analysis had different approaches for regulatory measures ^21^: while government decrees for social distancing policies were adopted from March 17^th^ in some capitals – especially in the South and Southeast – there was a considerable delay in others, noticeably in the North^21^. Thus, our analysis reflect different stages of the pandemic, and numbers may have a dynamic behavior over time. Moreover, the age structure of the studied cities, with greater proportion of older individuals in the Southeast, even emphasize the disparities in premature mortality in the less developed cities.

Our results, with a significant reduction of ACS and stroke in 4 capital cities, may look counterintuitive, considering the reported cardiovascular effects of COVID-19 that may occur through: direct myocardial invasion by the virus, increased metabolic demand, systemic inflammatory response contributing to an induced prothrombotic state, and also due to deleterious effects of empirical drug schemes precipitating arrhythmias^22-24^. Moreover, an increase in risk factors for cardiovascular disease - such as tobacco use and reduced physical activity - has also been reported during the pandemic^25^. From our data, thrombotic causes of death (ACS and stroke) increased only in the 2 Northern capitals experiencing healthcare collapse. However, the reduction observed in the other cities paralleled with the increasing occurrence of home deaths and deaths with unspecified cardiovascular cause. This may be explained by 3 factors: limited access to hospitals in locations where an overload was being experienced, avoidance of medical care due to social distancing or concerns of contracting COVID-19 in the hospital, and isolation that impairs the detection of cardiovascular symptoms by others^5 7^. The strong positive correlation between the rise in unspecified cardiovascular and home deaths corroborates these explanations, as it may suggest that at least some of the missed ACS and stroke deaths occurred at home, precluding correct diagnosis. Conversely, acute cardiovascular events may have decreased in some locations due to competing risks, and reduced exposure to secondary triggers of acute cardiovascular events, such air pollution^26^. More detailed insights will only be possible after the analysis of SIM.

Besides patient-based factors, reorganization of acute care systems, such as: deactivation of specific services to meet urgent needs of emergency or intensive care, delimitation of COVID-specific hospitals, and implementation of alternative therapeutic pathways; which aim to mitigate the effects of the pandemic, may further prevent patient presentation to medical care^4-7^. Thus, public campaigns must be warranted to raise awareness about the importance of cardiovascular care, even during this challenging period. Of note, the deleterious effects on cardiovascular events may last longer than the pandemic itself, as primary and secondary preventions are being delayed in this context^27^.

## Limitations & strengths

Our study has several limitations. At first, we utilized raw data extracted from the Civil Registry, without epidemiological adjustments. Thus, there was no investigation, codification or reclassification of deaths declared, nor a specific methodology for redistribution of garbage codes. As such, our data relies on how the DC are filled, what may differ across locations. Second, the data mining algorithm considered all causes reported in the DC, without hierarchical classification or identification of the underlying cause of death. This is a technical limitation of the ARPEN database, and might lead to misidentification of causes. Moreover, the delays in reporting may be differential across locations, and that is why capital cities – which are less prone to delays – were chosen for our analysis. Fourth, the availability of COVID-19 tests is heterogeneous between Brazilian regions, what may further impact case reporting. However, despite these limitations, to the best of our knowledge this is the most promptly available data source in Brazil on excess mortality in the COVID-19 pandemic, allowing for important epidemiological insights. The release of the SIM data, in the near future, will certainly provide more accurate estimates.

## Conclusion

In the 6 Brazilian capital cities with higher numbers of deaths due to COVID-19, there was an excess mortality, with greater magnitude in the more deprived cities regarding socioeconomic development and health resources. In general, cardiovascular deaths increased mainly as a result of unspecified cardiovascular causes, which correlated with the rise in home deaths, presumably as a result of impaired access to healthcare leading to misdiagnosis of specific cardiovascular causes, such as ACS and stroke. In the Northern cities, however, excess deaths occurred for ACS and stroke, in addition to unspecified cardiovascular causes, what differs from reports of HIC and is possibly associated with worse health infrastructure. As the pandemic advances to LMIC, investments in health resources, prioritizing socioeconomically disadvantaged locations, and public awareness campaigns focused on acute cardiovascular care are of utmost importance to mitigate the pandemic impact.

## Contributorship statement

Conception and design of the research: Lopes, MACQ, Oliveira, GMM, Ribeiro, ALP, Brant, LCC, Nascimento, BR; Acquisition of data: Teixeira, R, Lopes, MACQ, Oliveira, GMM, Malta, DC; Analysis and interpretation of data: Teixeira, R, Brant, LCC, Nascimento, BR, Ribeiro, ALP, Oliveira, GMM; Statistical analysis: Teixeira, R; Obtaining financing: N/A; Writing of the manuscript: Nascimento, BR, Brant, LCC; Critical revision of the manuscript for intellectual content: All authors; Authors responsible for the overall content as guarantors: Brant, LCC, Nascimento, BR, Ribeiro, ALP.

## Supplementary materials

**Supplement Figure 1:** Trends for deaths associated with acute coronary syndromes and unspecified cardiovascular causes in 6 selected Brazilian capital cities, in 2019 and 2020.

**Supplement Table 1:** Dictionary of search terms applied for the data mining procedure in the Civil Registry Database.

